# Digital contact-tracing during the Covid-19 pandemic: an analysis of newspaper coverage in Germany, Austria, and Switzerland

**DOI:** 10.1101/2020.10.22.20216788

**Authors:** Julia Amann, Joanna Sleigh, Effy Vayena

## Abstract

Governments around the globe have started to develop and deploy digital contact tracing apps to gain control over the spread of the novel coronavirus (Covid-19). The appropriateness and usefulness of these technologies as a containment measure have since sparked political and academic discussions globally. The present paper contributes to this debate through an exploration of how the national daily newspapers in Germany, Austria, and Switzerland reported on the development and adoption of digital contact-tracing apps during early and after stages of the lockdown. These countries were among the first in Europe to develop apps and were critical voices in the debate of decentralized vs. centralized data processing. We conducted thematic analysis on news coverage published between January and May 2020 in high-circulation national daily newspapers (print) from Germany, Austria, and Switzerland. A total of 148 newspaper articles were included in the final analysis. From our analysis emerged six core themes of the development and adoption of digital contact tracing apps: 1) data governance; 2) role of IT giants; 3) scientific rigor; 4) voluntariness; 5) functional efficacy; 6) role of the app. These results shed light on the different facets of discussion regarding digital contact tracing as portrayed in German-speaking media. As news coverage can serve as a proxy for public perception, this study complements emerging survey data on public perceptions of digital contact tracing apps by identifying potential issues of public concern.

## Introduction

### Covid19 outbreak and pandemic

The novel coronavirus (Covid-19), first detected late 2019 in China, quickly reached global proportions, leading the World Health Organization (WHO) to declare a pandemic on March 11, 2020. Researchers found the virus to be highly contagious with cases ranging from asymptomatic to severe respiratory infections.[1] To control the outbreak, several countries went into lockdown, an approach that restricted individual freedom of movement, hurt the economy, and abruptly halted international travel. While such draconic measures succeeded in containing the virus and reducing transmission rates [2, 3], their full economic and societal impact remains unclear. As governments then began to restart economic and social life post lockdown, the need grew for suitable exit strategies to prevent a second wave of infections that would overwhelm healthcare systems.[3-5] To this end, many lands and states introduced several non-pharmaceutical interventions, including social distancing and hygiene policies. Among such interventions, the identification and isolation of positive cases through contact tracing plays an instrumental role.[6]

### Digital contact tracing

Since the Covid-19 outbreak began, digital contact tracing’s usefulness as a containment measure has sparked political discussions globally.[7] As one of the oldest public health disease methods, contact tracing works by public health experts identifying infected individuals, isolating them and then finding out with whom they came in contact. Digital contact tracing expands on this by harnessing mobile technologies such as GPS, Bluetooth or QR codes, to digitally track and notify users about their interactions with potentially infected individuals.[8] This automated and digital approach thus offers governments a more cost-effective and easily scalable method than traditional contact tracing.[9]

Given such advantages, several countries have or are in the process of developing Covid-19 digital contact tracing mobile applications. Asian countries with previous experience of SARS, in particular China and Singapore, quickly took on a pioneering role, capitalizing on their well-developed digital infrastructures to deploy contact tracing apps amongst other digital disease surveillance methods. Concurrently, researchers in Europe formed collaborations to work on contact-tracing solutions, with one of the first being the Pan-European Privacy-Preserving Proximity Tracing (PEPP-PT) consortium. Austria was then among the first countries in Europe to deploy a digital contact-tracing app. Their “Stopp Corona-App”, launched by the Austrian Red Cross, was available for download from March 25, 2020. In Germany, the Robert Koch Institute launched its “Corona-Warn-App” on June 16, 2020. Switzerland’s “SwissCovid-App”, jointly developed by the two Swiss federal institutes of technology (EPFL and ETH), then followed on June 25th. These German speaking countries use digital contact tracing as a tool solely to support mitigation and containment, contrasting to other countries (like Taiwan, Singapore and Poland) that use digital contact tracing apps to enforce quarantine measures.

### Challenges and opportunities of digital contact tracing

The academic community rapidly responded to this surge in digital contact-tracing solutions. Critics argued that such apps bring to the fore several ethical and technical challenges. To begin, scholars questioned the approach’s accuracy. Experts highlighted the low data quality and Bluetooth technology’s inaccuracy for proximity tracing, and that including citizen’s self-diagnosis along with official validated tests propagates false positives.[7, 10] Furthermore, although a significant percent of the population needs to participate for efficacy, in countries like Singapore the adoption rate has been below 20%.[11] The opt-in approach can thus undermine effectiveness through lack of critical mass, however making it mandatory would sacrifice autonomy.[12, 13] Furthermore, there are short and long-term surveillance and privacy concerns.[8] Critics highlight that digital contact tracing operationalizes a large-scale surveillance system that could outlive the pandemic.[14] Others draw attention to potential privacy breaches, the questionable reliability of anonymization, the national differences in data protection and privacy regulations, as well as the weaknesses of centralized report processing.[15, 16] Despite this myriad of concerns and challenges, proponents maintain that digital contact tracing, when combined with other measures, careful oversight, and abidance to data protection regulations and protocols through privacy preserving technological options, can reduce transmission rates and allow for a relaxation of measures.[13]

### Public perceptions of digital contact tracing

Despite a rapidly growing body of official recommendations and academic literature, to date, evidence on end-users perceptions and adoption behavior of digital contact tracing apps is only starting to emerge.[17, 18] Many studies on the topics use online surveys to collect data on public perceptions. For example, one survey of the Swiss public in April and May found that almost three-quarters of the Swiss population (72%) said that they would be willing to install such an app if it could help slow down the spread of the corona virus and shorten the length of the lockdown.[19] Yet, a July survey of the Swiss public indicated that only 1 out of 3 citizens downloaded the SwissCovid app, with the main reasons against being not having a suitable smartphone (28%), perceiving it as not useful (26%) and privacy concerns (24%).[12] These studies start to reveal differences between declared download intentions and actual download and usage behavior, a phenomenon known as the intention-behavior gap.[20] However, while several surveys are currently underway, to date, evidence on public perceptions and usage intentions remains scarce and inconclusive.

Another way to explore public views on digital contact tracing apps is by looking at how the media frame these technologies. News media represent an important source of information through which the public learns about health issues and new technologies. Despite the decline in print and move towards digitalization, newspapers still serve as useful proxies for reporting across other channels.[21] Evidenced by earlier work, news media can shape public perceptions of a given topic in different ways, namely through: the amount of coverage of a particular topic during a given time; the specific content of media messages; and the tone of coverage.[22] In the field of crisis and risk communication, research also shows that news coverage influences public risk perception and can impact community health behaviors and practices.[23-26]

### Study Aims

Building on this literature this study aims to investigate how national daily newspapers in Germany, Austria, and Switzerland report on the development and adoption of digital contact-tracing apps during early and after stages of the lockdown. Findings of this study are timely and can inform the rapidly evolving technical and political work on digital contact tracing apps. By understanding how the media frame digital contact-tracing apps, government and health departments can more strategically communicate with the public. The identification of what information and issues are most prevalent in the media can further help to direct efforts that address public concerns to support public acceptance and uptake of digital contact tracing apps.

## Materials and Methods

We conducted a thematic analysis [27] of newspaper articles from Germany, Austria, and Switzerland published between January and May 2020 reporting on the development and adoption of smartphone apps for digital contact-tracing during the Covid-19 pandemic. In contrast to qualitative content analysis, which uses a descriptive approach to coding and interpreting quantitative counts of the codes, thematic analysis seeks to present a purely qualitative, detailed, and nuanced account of the data analyzed.[28]

### Data collection

For this study, we focused on news coverage of digital contact-tracing apps in high-circulation national daily newspapers (print) from Germany, Austria, and Switzerland. We chose to focus on these countries, as they were among the first in Europe to develop digital contact tracing apps, and were key voices in the debate of decentralized vs. centralized data processing. In our selection, we included broadsheet and tabloid press to cover a wide sample of newspapers with various readership profiles.[29] We retrieved newspaper articles from the Factiva global news monitoring and search engine on May 29, 2020, using the search strategy displayed in Textbox 1. We chose to focus our search on the first months of the pandemic (Jan-May 2020) to cover periods pre-lockdown, during lockdown, and the beginnings of post lockdown. The search yielded a total of 459 news articles. As part of an initial full-text screening, we filtered out duplicates and news items that did not, or, only briefly mentioned digital contact-tracing apps.

**Textbox 1. Search strategy**

- **Time period:** 01.01.2020-29.05.2020
- **Search terms:** “contact-tracing” OR “corona-app” OR “proximity-tracing”
- **Language:** German
- **Source:** national daily newspapers (print) in Germany, Austria, and Switzerland
  ∘ **D:** Süddeutsche Zeitung, Frankfurter Allgemeine Zeitung, Die Welt, Bild
  ∘ **A:** Der Standard, Die Presse, Kurier (Krone Zeitung: not available)
  ∘ **CH:** Tages Anzeiger, NZZ, Blick

### Data analysis

Eligibility screening led to the inclusion of 153 news articles which we imported into MAXQDA (VERBI Software, 2019), a qualitative data analysis software. After an additional round of full-text screening, 5 articles were removed because they did not fully meet the inclusion criteria. A total of 148 news articles were included in the final analysis.

To develop a draft coding scheme, the research team first deductively identified content areas of interest based on digital contact tracing’s political and academic discourse. In a first coding phase, the researchers extended this preliminary codebook through an additional round of full-text screening, following an inductive approach. During this stage, the researchers used memos to document coding category descriptions and instructions about their use.[30] In the next step, two researchers (JA, JS) independently applied the preliminary codebook to a sub-sample of the articles (N=30). During this process, new coding categories were added, and others merged, modified, or removed. Codes were assigned to text written directly by the journalist as well as quotes from interviews. The two researchers iteratively compared their coding and refined the codebook. The refined codebook was then again applied to a sub-sample of the articles (N=10) by both researchers independently. Intercoder reliability corrected for agreement by chance (Cohen’s Kappa) indicated an average reliability of 0.97. One researcher (JA) coded the remaining articles. During regular meetings, we finally gathered and collated codes into overarching themes in an iterative process guided by the study aim. Exemplary extracts are provided to illustrate our findings.

## Results

In the search period (Jan-May 2020), we identified a total of 148 articles that focused on digital contact tracing. These were published between March 19 and May 29, 2020 and came from nine newspapers. The majority of articles (69 articles, 46.6%) were Austrian, from the newspapers: Der Standard (17 articles), Die Presse (30 articles), and Kurier (22 articles). Germany had 26 articles (17.6%) from Bild (2 articles), Die Welt (13 articles), and the Süddeutsche Zeitung (11 articles). From Switzerland were 53 articles (35.6% articles) from Blick (12 articles), Neue Zürcher Zeitung (23 articles), and Tages Anzeiger (18 articles). Fig 1 shows the distribution of articles over time alongside key events in each region during the investigated period. Note, Fig 1 only reports Austria’s launch of a contact tracing app, as Germany and Switzerland had not released their apps during the time this research was carried out.

**Fig 1.**
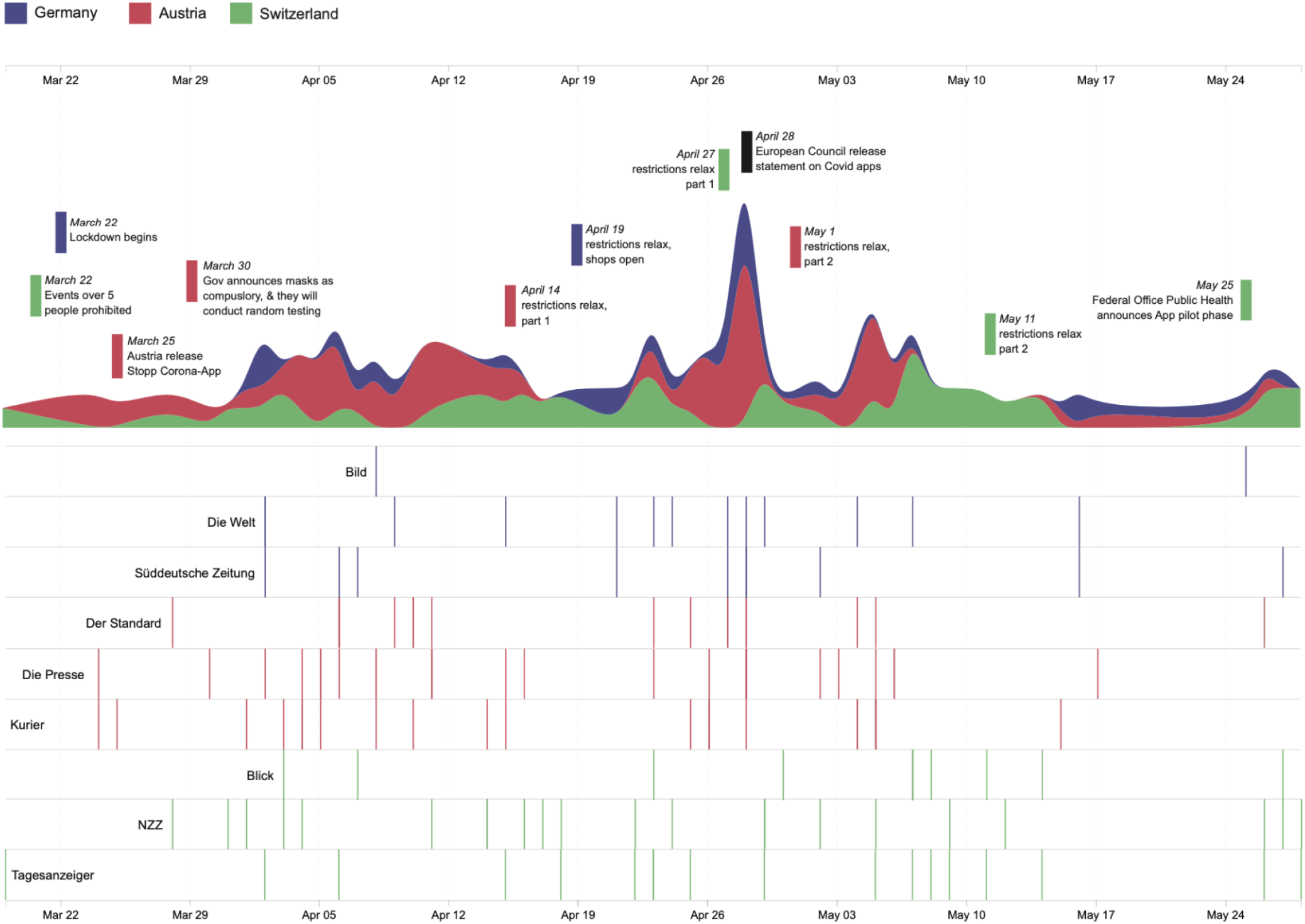
Distribution of analyzed articles published between March 19 and May 29, 2020. Colors represent regions represented. Important events are also plotted.

Based on our analysis, we identified six core themes in the news coverage on the development and adoption of digital contact tracing apps: 1) data governance; 2) role of IT giants; 3) scientific rigor; 4) voluntariness; 5) functional efficacy; 6) role of the app. An overview of the themes is presented in Fig 2. As well, we observed no apparent regional differences in the occurrence of themes in the news coverage. Fig 3, however, highlights how discussions of app development and app adoption went hand in hand, with app adoption themes slightly more present.

**Fig 2.**
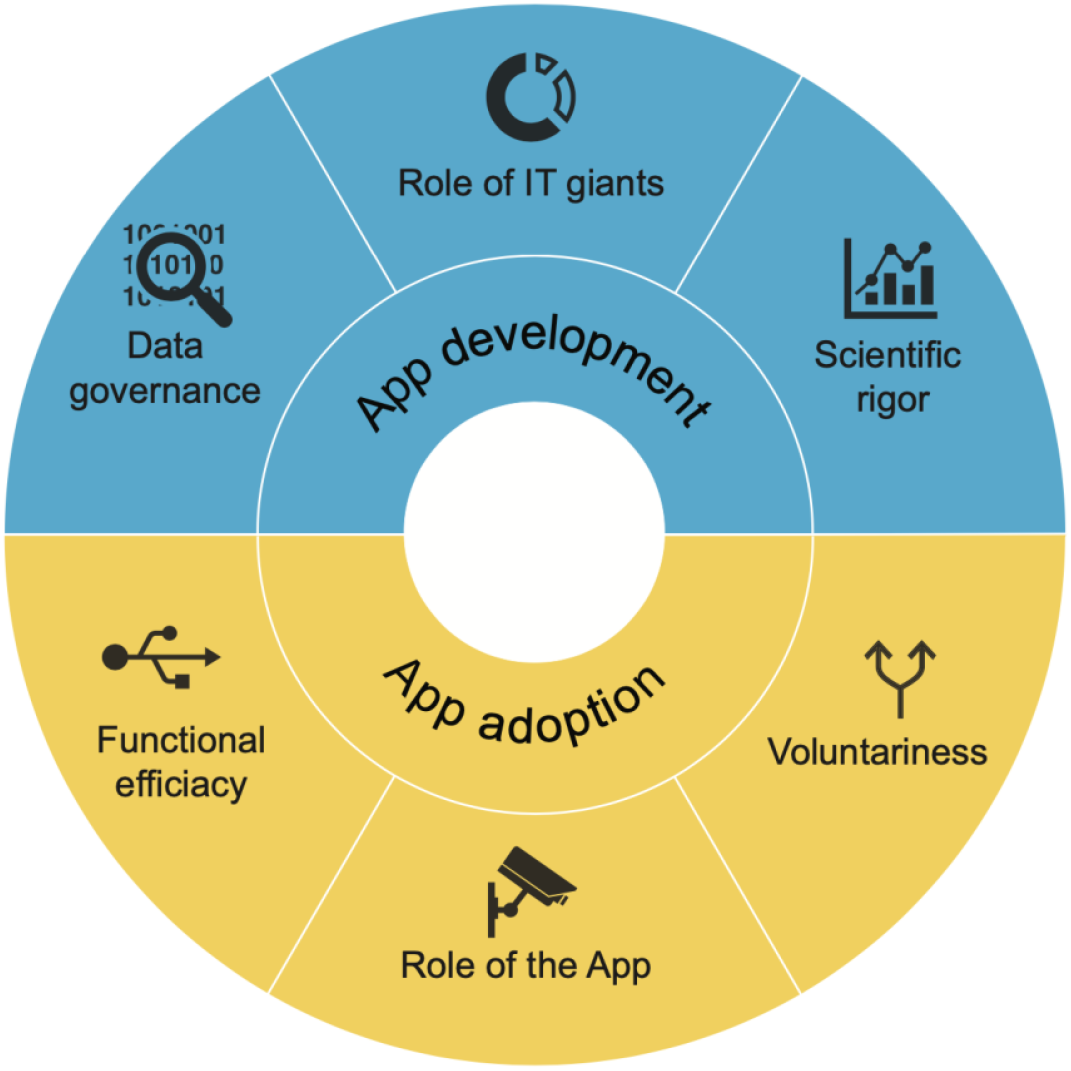
Results of the thematic analysis.

**Fig 3.**
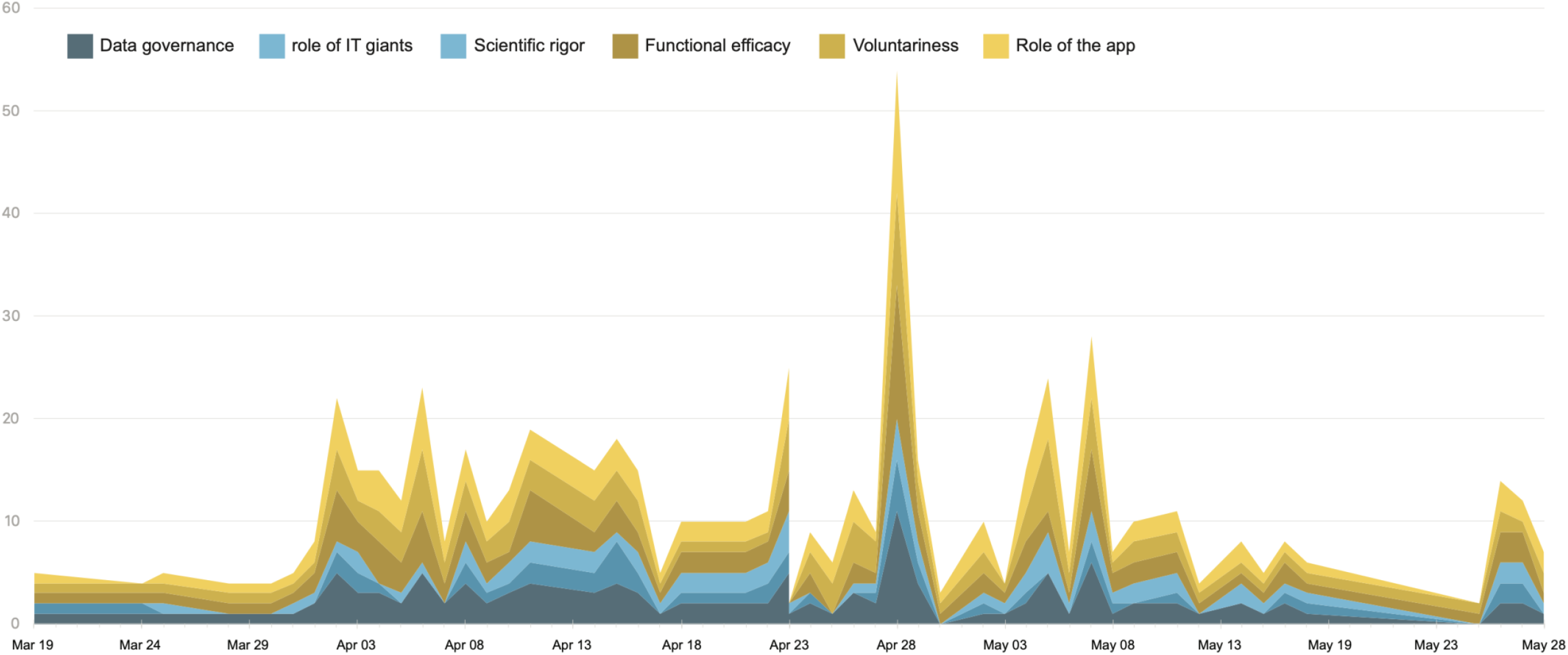
Theme occurrences per article over time.

### App development

#### Data Governance

Data governance was a central theme in news coverage on digital contact tracing apps. It encompasses discussions surrounding the collection, storage, and access to data generated by digital contact tracing apps, covering also related issues of data protection, privacy, and anonymity. A primary point of debate was whether apps should store data centrally or decentralized, with arguments for both approaches being acknowledged. One article indicated that with a centralized approach “*only an anonymized code is centrally recorded from infected persons*” (Tages Anzeiger, 11 May) and that the aggregated data might help authorities “*to better understand the virus and its spread and the effectiveness of their containment measures*.” (NZZ, 17 April). Several other articles, in contrast, warned against central solutions for reasons of data protection. One article explained, “*even if the technology only stores anonymous numbers instead of names, a reconstruction or de-anonymization would be possible based on the data*.” (Die Welt, 21 April). Bolstering support for decentralized approaches was then an open letter from 300 researchers pleading not to *“create a tool that would make it possible, now or in the future, to collect data on the population on a large scale,”* (Tages Anzeiger, 22 April). This letter marked the escalation of a dispute amongst the Pan-European Privacy-Preserving Proximity Tracing (Pepp-PT) consortium.

Linked to data governance, data protection was another key subtheme. To start, some articles detailed how, unless carefully developed, contact tracing apps would undermine privacy, meaning the ability to control who can access or use ones’ data. Several articles thus called for ‘privacy by design’, reasoning that “*we are not a flock of sheep, but lead our own lives, red lines must not be crossed in tracking tools*.” *(Die Presse, 28 April)*. Even though one article indicated that *“experts largely agree on the criteria an app and its underlying software must meet to ensure that data and privacy are protected”* (NZZ, 14 April), the dispute around centralized vs. decentralized approaches suggests otherwise.

Articles also stressed the importance of ensuring anonymity, meaning *“that the data is secure and anonymous and that no one, especially not the state, will access it*.*”* (Die Presse, 3 May). In regards to the Austrian app, one article explained that to protect privacy, an integrated statistics function was removed *“because it would have allowed users to be identified and traced*.*”* (Die Presse, 17 May). Also in Germany, articles discussed how exactly data would be collected “*solely to let users know whether they have been in close contact with other, already infected users - without revealing their identity”* (Süddeutsche Zeitung, 16 May). Several articles went further into technical details regarding how anonymized IDs and decentralized systems could mask user identities (e.g. NZZ, 18 April).

Articles also contextualized privacy trade-offs in terms of western vs. eastern values and cultures. Some articles used Eastern countries as warning examples - “*in contrast to tracing apps, such as those used in Asian countries, the privacy of users should therefore be protected according to European standards*.*”* (NZZ, 16 April). South Korea’s *“surveillance state*” approach (NZZ, 28 March), and China’s extensive data collection (Süddeutsche Zeitung, 16 May) repeatedly came up in such discussions.

#### Role of IT giants

The theme “Role of IT giants” covered aspects related to the role of technology companies in the app development phase. When reporting on the role of IT giants, Apple and Google were most frequently named. On the one hand, several articles portrayed these companies as supportive collaborators who joined forces to help governments and the global community fight covid-19 through the development of contact tracing technologies. “*The companies are working together to pave the way for Corona apps on the software side*.*”* (Die Presse, 28 April).

On the other hand, many articles portrayed the companies as dominant and powerful decision-makers, questioning Europe’s dependency on US corporations and infrastructures. For example, one article reported that the German government’s pivot from a centralised to decentralized approach was motivated mainly because Google and Apple did not support a centralized approach. Another article went so far as to say that the big IT players would *“force governments to save and analyze corona data in a certain way”* thereby “*disempowering democratically legitimized decision makers*” (Süddeutsche Zeitung, May 27). Highlighted was also how the big tech players were gatekeepers that *“dominate the market for mobile operating systems”* (Süddeutsche Zeitung, April 27). As one article stated *“the app will only achieve wide dissemination once it is possible to download it in the Google and Apple app stores* (Blick, May 8).

In addition to power dynamic concerns, some articles questioned Apple & Google’s trustworthiness and commitment to data protection. One article emphasized that “*these companies are not democratically legitimized”* and that *“they have often proven that they care about general public interest as little as they care about European laws”* (Die Welt, April 28). Another article quoted an Austrian politician voicing concerns that *“data might flow to giant companies like Google and Microsoft*.*”* (Kurier, April 15).

#### Scientific Rigor

The theme “Scientific rigor” encompasses those instances where articles discussed issues relating to transparency and testing in the app development process. Several articles discussed the importance of disclosing the source code, *“the holy grail of a program”* (Die Presse, May 3). Many of the articles asserted that this disclosure represents an important control mechanism and that only code subjected to public scrutiny could be officially recommended. As one article put it: *“That means that the source code of the program is publicly accessible, so that no one can implement a loophole in the software unnoticed”* (Tages Anzeiger, April 22). In this context, newspapers also referred to audits by external experts, including hacker communities and data protection specialists.

Another aspect was communication with the public. In some articles, governments or politicians were criticized for their lack of transparent communication about the purpose and functioning of contact tracing apps. One article, for instance, highlighted the need to go beyond conventional *“marketing efforts to calm down the public”* and to instead explain *“in detail what data is needed and how the Stopp-Corona-App system works”* (Die Presse, April 28). Similarly, other articles emphasized the need to be transparent *“about the integration of measures into the overall strategy”* and to *“inform and clearly communicate when and under which circumstances they will no longer be used” (NZZ, May 5)*.

Pertinent to the theme of Scientific Rigor, we also identified several articles reporting on digital contact tracing apps’ pilot or testing phases. In contrast to the disclosure of source code and communication to the public, pilot phase reports were mostly neutral. In this sense, articles mainly outlined the setting, and who was involved in pilot-testing. Only a few articles explicitly stated the pilot phase’s purpose, acknowledging that it *“can be used to detect potential flaws of the app and to strengthen its acceptance”* (Blick, May 7).

### App adoption

#### Voluntariness

Voluntariness was a recurring theme of app adoption in our analysis. Several articles addressed the question of whether digital contact tracing apps should be voluntary or mandatory. Voluntariness was stressed as *“an important point when it comes to being in line with data protection regulations”* (NZZ, April 16). Yet, other articles queried whether a voluntary app could achieve a wide enough adoption to fulfill its purpose, leading to questions like: *“Will the corona-app become mandatory?”* (Blick, April 23), “*Can it work out with voluntariness?*” (Die Presse, April 28), *“Is participation voluntary?”* (Die Welt, May 7). In this context, the issue of proportionality was raised. One article quoted an Austrian data protection expert stating that a mandatory app would be possible *“if the alternative would be the complete prohibition of many activities”* meaning *“the corona-app would represent the lesser of two evils”* (Die Presse, April 28). This view was echoed in a Swiss news article, which outlined the benefits of a mandatory approach, describing it as an “*alternative to a lockdown*” whilst acknowledging that *“the legal basis is not clarified”* (Tages Anzeiger, April 23). In contrast, another Swiss article quoted a Swiss data protection officer stating that voluntariness was non-negotiable and that *“an obligation would be an unproportionate infringement of citizen’s self-determination”* (NZZ, May 2).

Unsurprisingly, the discourse surrounding the question of voluntariness also evoked political debate, with parties voicing concerns that the app *“would de facto become an entry ticket to events”* and thereby create *“an indirect obligation to use the app, at least for those that do not want to be excluded”* (Der Standard, April 27). The Austrian government’s considerations to make the app mandatory led to outrage from the opposition who warned that the app would become an *“electronic ankle monitoring device”* (Der Standard, April 6). Similarly, other articles reported political concerns that even if proclaimed voluntary, the app could still be *“forced based on economic and moral grounds”* (Blick, April 30) or subject individuals to *“a great degree of social pressure”* (Die Welt, April 2). These articles hinted at the risk of pressuring conformity, “*a soft obligation*” (NZZ, April 16), and thereby discrimination for those unwilling or unable to use the app. As in the case of data protection and privacy issues, articles often cited warning examples of other particularly Asian countries where app usage was mandatory.

#### Functional efficacy

The theme “functional efficacy” captured descriptions on how digital contact tracing apps function, their technological limitations, and societal aspects related to app download and usage as prerequisites for successful deployment. Specifically, articles explained in various levels of detail the technological and logistical workings of app-based contact tracing. For example, while some articles precisely reported how the apps measured duration and distance, others would simply refer to *“the use of Bluetooth technology to determine which people were in close contact”* (Tages Anzeiger, May 11). Regarding logistics, some articles explained what users should do if tested positive or receiving the app’s contact-warning message. However, the information provided wasn’t always clear, with many questionings: *“What happens after receiving a warning via the app?”* (Die Welt, April 21). Some articles quoted politicians’ suggestions to favor app users for virus testing and to incentivize quarantine to users who receive warnings, yet, no information was provided on how such a system would or could be implemented. In an interview, a Swiss epidemiologist acknowledged that *“there is a need for clearly defined processes which still need to be developed”* (Tages Anzeiger, April 15). Even when the Austrian app was already available, open questions remained, such as *“What do I do if someone gives notification about having corona symptoms but did not test yet?”* (Die Presse, April 28).

Within the theme of functional efficacy, we also identified articles that acknowledged the technological limitations of Bluetooth based apps (the designated technology used in all three countries). These articles raised important questions as to how to ensure effectiveness while minimizing the risk of false positives and negatives. One article, for example, explained there are *“hundreds of different smartphones with technically different Bluetooth chips, antennas with different sending characteristics, smartphone cases with different levels of damping”* (Die Welt, May 7). According to the article, these factors, and that smartphones might be in a bag could lead to exposure risk underestimation. On the other hand, another article suggested the overestimation of exposure risk because *“the app can only measure distances, thus also detecting contacts in cases where there is only little risk of infection due to protective measures, like face masks or glass panels”* (NZZ, May 26). Also, the question of interoperability between different national digital contact tracing apps remained unanswered.

Two frequently addressed aspects were app download and usage intentions. Here, several articles stated that correct app usage was critical to success and simultaneously the greatest hurdle. As one article wrote: *“The app only makes sense, if it’s used correctly by as many Austrians as possible”* (Kurier, March 24). While some articles reported the actual app download numbers and users, others provided information on the required download and usage rates for efficacy. In terms of the needed level of adoption, several articles provided unspecific numbers, such as *“as many people as possible”* (Süddeutsche Zeitung, April 28), *“millions of people”* (NZZ, April 16), or *“enough people”* (Die Welt, April 2). Others instead, reported concrete percentages ranging from 20% to 60% of required adoption rates. However, for the most part, it remained unclear how and by whom these estimates were derived.

Articles also reported surveys on public acceptance and download intentions (e.g. NZZ, May 26; Bild, May 25; Tages Anzeiger, May 5), and speculated factors promoting or impeding app adoption. One article, for instance, noted that *“centralized data storage could become a problem as it would most likely undermine public trust and acceptance of the corona-app”* (Die Welt, April 21). Another questioned whether digital contact tracing apps could be successful *“in a county where people worry more about data protection than in Singapore”* (Die Welt, April 24). Yet, other articles were more optimistic. One article quoted a Swiss politician: *“It is about the wider public. They have adhered to the rules in the past weeks - they will also use the app*.*”* (Blick, May 11). The need of fostering public trust was also frequently mentioned in relation to public acceptance. Aptly, one article noted that “*a question arises not only technically, but also politically and socially: Who do we trust?”* (Die Welt, 28 April). Others acknowledged that “*we are giving the government more and more power to monitor us*.*”* (Kurier, 15 May) *and that “without the trust of citizens voluntary apps will not work”* (NZZ, April 17).

#### Role of the app

The theme “role of the app” encompasses those instances where articles referred to the potential role digital contact tracing apps could take on. Although the media’s portrayal of digital contact tracing apps ranged between positive/optimistic to negative/pessimistic, the majority of articles took a positive stance. Whereby articles emphasized digital contact tracing apps as a necessary means to return to normality whilst *“avoiding a second wave”* (Blick, May 7). They described digital contact tracing apps as *“digital helpers that should play a decisive role on the way back to normality after the coronavirus pandemic”* (NZZ, April 17) or as a “*necessity for the slow exit from the lockdown*” (Die Welt, April 21).

Digital contact tracing apps were also frequently depicted as an important tool that *“can warn individuals when they have had contact with someone who later turns out to be infected”* (Blick, April 30). Similarly, articles reported apps as a means to gain control over the virus i.e. *“keeping reproduction number of the virus below 1”* (Tages Anzeiger, May 26); *“to slow down the spread of the virus*” (Süddeutsche Zeitung, April 28); *“to break the chain of infection”* (Kurier, March 25). Some articles also portrayed the app as a tool to foster self-determination, responsibility, and solidarity, referring to it as *“an instrument for exercising self-determination and personal responsibility*.*”* (NZZ, May 28).

Despite the predominantly positive coverage, several articles also acknowledged that digital contact tracing apps are “*not a panacea but at best one of many building blocks in the fight against the virus”* (Die Welt, May 27). Along these lines, articles emphasized that digital contact tracing apps *“will not stop the coronavirus on their own*.*”* (Der Standard, April 10) and that *“in the fight against Covid-19, technical solutions are not the most important thing but people are”* (Die Welt, April 24). As such, digital contact tracing was portrayed as *“one instrument of many”* (Die Presse, April 28) in the fight against the virus. Articles particularly emphasized its complementary role to traditional contact tracing: *“The planned app is not a replacement but ‘just’ an addition to conventional contact tracing”* (NZZ, May 27).

Finally, there were also articles highlighting the potential dangers of apps as a “*low-threshold entry into a nasty monitoring tool that records movement profiles*” (Süddeutsche Zeitung, April 28) leading to the “*installation of a surveillance state*” (Kurier, April 15). One article even referred to the digital contact tracing practices of autocratic regimes as warning examples that *“hollow out their citizens’ privacy via smartphone in favor of the common good”*, yet acknowledging that also *“western democracies are preparing to have direct access to data collected via smartphones, in case of emergency” (NZZ, April 14)*.

## Discussion

Scientific evidence, political debates, and media coverage surrounding digital contact tracing apps continuously evolve in real time and shape public perceptions. In recognition of this flux, this work documents how in the early stages of the pandemic national daily newspapers in Germany, Austria, and Switzerland reported on the development and adoption of digital contact-tracing apps. Being among the first countries in Europe to develop and deploy digital contact tracing apps, these three German-speaking countries took on a pioneering role and were key voices in the centralized vs decentralized debate. The news media coverage in these countries thus presented a particularly rich setting to study. While we did not assess public response to news coverage, findings of this study can complement emerging survey data on public perceptions of digital contact tracing apps by identifying potential issues of public concern.

Our findings indicate that the media discourse was characterized by conflicting views on the role and modalities of digital contact tracing apps. In particular, debates surrounding data protection, privacy, and voluntariness were featured prominently in our sample, reflecting also the academic discourse.[12] When expert controversies occur, the public relies on the authority of researchers and health professionals as communicated in the media, yet, the difficulty of judging competing expert claims leads to confusion, for if experts cannot agree, who should the public trust and follow?[31] Research has shown that conflicting messages can create skepticism and lead to decreasing public trust in governments’ ability to manage a public health crisis.[32] Moreover, presenting conflicting expert statements or contradictory data can decrease an individual’s adherence to recommended measures.[33, 34]. The media’s presentation of digital contact tracing apps as controversial could have, therefore, led the public to be cautious and reluctant to use the app.

Interestingly, the results of our analysis also mirror a growing resistance to governmental measures and restrictions as demonstrated by Covid-19 protests in European cities.[35] This anti-establishment sentiment emerged, to a certain extent, in the articles analyzed. For example, many articles raised questions of whether authorities could be trusted to uphold data protection and privacy. As well, several articles raised doubts about the trustworthiness of Google and Apple as dominating US players in contact tracing app development. Discussing Europe’s dependency on US infrastructures, along with the questionable data protection practices of these companies could have also spurred mistrust in governmental efforts. In Switzerland, however, this doesn’t seem to be the case. While a representative survey carried out at the end of June 2020 indicated that the majority of the Swiss public would not be willing to install the app mainly due to concerns of data protection,[36] findings of more recent surveys paint a different picture. One survey conducted by the University of Zurich, for example, showed that more than half of the respondents (54%) were willing to download a digital contact tracing app if it were launched by the Swiss Federal Council, about a third (33%) would do so if the app were launched by the Swiss Federal office of Public Health, 28% if the publisher was a cantonal government. Fewer respondents would install an app launched by other organizations (e.g. universities, or international / non-profit organizations) or technology companies.[19] Findings of the survey further indicated that limiting the spread of the virus and shortening the duration of the lockdown were main drivers for willingness to download the app.[19] In line with these findings, a national representative survey conducted at the end of July found that solidarity was the most important reason for activating the SwissCovid App.[37] Findings of a German study similarly suggest that app users perceive the benefits of using the app outweigh potential risks, and they trust that the app can help to slow down the pandemic.[38] These findings mirror the results of our analysis, which showed that newspapers adopted a predominantly positive stance towards digital contact tracing apps, highlighting their role to gain control over the virus and facilitate a return to normality.

A recurrent theme in our analysis related to functional efficacy and the potential technical and logistical challenges of digital contact tracing apps. Raising awareness to these issues may have led the public to question the overall effectiveness and usefulness of these technologies in the fight against the pandemic. Yet, contrary to these issues of functional efficacy we identified in our analysis, findings of a Swiss national representative survey indicate that app users are satisfied with the app, judging the installation and functioning of the app as simple and comprehensible.[37] Only few respondents reported error messages or unclear notifications. This may indicate that some of the initial problems that were covered by the news media were resolved. At the time of writing this article, the SwissCovid-App Monitoring website reported a total of 2’373’287 app downloads and a fairly steady number of active users ranging between 1’630’000 and 1’600’000 in the period from Sept 9 - Sept 15, 2020.[39] Even though not all downloads convert into active app users, the steady number of active users does indicate at least some public support. In contrast to these survey results, there are emerging media reports on technical and logistic issues for the digital contact tracing apps.[40-42]

According to recent numbers published by the Robert-Koch-Institute, the German Corona-Warn-App counted 18.2 Mio downloads (as of Sept 15 2020) and an average 1’390 calls to the Corona-Warn-App hotline in the period from Sept 7 - Sept 14, 2020.[43] While no official data on active app users is available for the Austrian Stopp-Corona-App,[44] findings of a recently published marketing survey suggest that uptake of the app remains lower in Austria compared to Germany and Switzerland.[45] Overall, these numbers indicate that app adoption rates remain low - a concern that was already identified by the news media in the early stages of the pandemic, as shown by our analysis. Thinking of individuals as “conditional contributors” to the public good, we would expect that they partake in activities they see others partaking in.[46, 47] This would mean that if the public is given the impression that only few people are following recommendations to use the app, they may find it fair to also refrain from using the app themselves. Based on this, the publication of low or stagnating app adoption rates could undermine the contact tracing app approach.

### Limitations

One of the limitations of this study was that the time it was carried out, only Austria had introduced a digital contact tracing app. This might explain why the majority of included articles were Austrian. Extending the period of data collection may have resulted in a more even distribution of samples. Despite this limitation, our analysis captured a wide breadth of themes that can serve as a basis for identifying public concerns of digital contact tracing apps. Another limitation was that we focused on daily print newspapers as opposed to popular online sources of information like social media. However, as outlined by earlier work, newspapers remain an important source of information that can serve as a proxy for reporting across other channels.[21] Meaning, that although our findings may not comprehensively portray the public discourse, they provide a useful overview of how news media depict digital contact tracing apps.

## Conclusion

To be successful, governmental measures, like digital contact tracing apps, need to be accepted and actively supported by the wider public. Yet, as with other controversial topics, like vaccination, and as evidenced by the growing number of demonstrations against governmental measures, achieving public consensus on digital contact tracing apps currently seems unlikely. To foster public trust and acceptance authorities thus need to develop clear and coherent communication strategies that listens to and addresses public concerns. In times of uncertainty, with new and sometimes contradicting evidence emerging almost on a daily basis, this is, however, not an easy task. As news reporting can serve as a proxy to public perception, findings of this study in complementation to emerging survey data can serve as a basis for identifying public concerns.

## Data Availability

Relevant data (translated text excerpts from news paper articles) are presented within the manuscript. The full data set (newspaper articles) cannot be shared due to copyright restrictions from the respective publishers.

